# An Overlooked Signal on Routine Bone Scans: Soft Tissue Uptake, Mortality and Cardiovascular-Kidney-Metabolic Burden

**DOI:** 10.64898/2026.06.08.26355179

**Authors:** Clemens P. Spielvogel, Kilian Kluge, Shaun Khanna, Tara Zuhair Kassem, Jing Ning, Katarina Kumpf, Christian Nitsche, Christian Hengstenberg, Piotr J. Slomka, Marcus Hacker

## Abstract

**IMPORTANCE:** Bone scintigraphy is among the most widely performed nuclear imaging examinations, with use in oncology, rheumatology, orthopedics and cardiology. Scans without cardiac uptake are read as normal with regard to cardiac amyloidosis and their soft tissue signal is dismissed as background. Whether this discarded signal is prognostic is unknown.

**OBJECTIVE:** To determine whether the soft tissue-to-bone ratio (STBR) on routine bone scintigraphy is associated with outcomes and cardiovascular-kidney-metabolic (CKM) burden.

**DESIGN, SETTING, AND PARTICIPANTS:** This retrospective cohort study included consecutive patients without cardiac uptake who underwent whole-body bone scintigraphy for any indication at a tertiary care center in Austria from January 2010 to August 2020.

**EXPOSURE:** STBR measured by blinded observers (continuous and at prespecified threshold, 0.5).

**MAIN OUTCOMES AND MEASURES:** The primary outcome was all-cause mortality; secondary outcomes were major adverse cardiovascular events (MACE) and heart failure hospitalization. Cox models were adjusted for 10 risk factors.

**RESULTS:** Among 8,769 patients (mean [SD] age, 61 [16] years; 3,066 [35%] male), 2,418 (28%) died during a median (IQR) follow-up of 5.1 (2.5-8.2) years. STBR greater than 0.5 (772 [9%]) was associated with all-cause mortality (adjusted hazard ratio [aHR], 1.73; 95% CI, 1.54-1.94), MACE (aHR, 1.51; 95% CI, 1.11-2.05) and heart failure hospitalization (aHR, 1.31; 95% CI, 1.02-1.67), with an increasing risk gradient. STBR per SD was associated with mortality in both sexes and every prespecified subgroup, including patients younger than 65 years (aHR, 1.25; 95% CI, 1.18-1.32), without cancer (aHR, 1.45; 95% CI, 1.28-1.64) and without renal insufficiency (aHR, 1.23; 95% CI, 1.18-1.28). Across 587 evaluated clinical traits, STBR mapped to cardiac, renal, metabolic, hepatic and inflammatory disease. Mean STBR increased from 0.32 at CKM stage 0 to 0.41 at stage 4 (ρ=0.30; P<0.001), strongest in patients without cancer (ρ=0.40; P<0.001).

**CONCLUSIONS AND RELEVANCE:** In this cohort study of patients with and without cancer or kidney disease, a signal dismissed as background on bone scans read as normal was independently associated with mortality and cardiovascular events while tracking CKM burden. If externally validated, STBR could enable opportunistic screening and risk stratification from bone scans without additional imaging, radiation or cost.

**Key Points:** *Question:* Does soft tissue signal on bone scintigraphy carry prognostic information in patients without cardiac uptake?

*Findings:* In this cohort study of 8,769 patients without cardiac tracer uptake, a higher soft tissue-to-bone ratio was significantly associated with all-cause mortality, major adverse cardiovascular events and heart failure hospitalization after adjustment for 10 risk factors, and in all prespecified subgroups. The ratio increased significantly across cardiovascular-kidney-metabolic stages.

*Meaning:* Soft tissue-to-bone ratio from routinely acquired bone scans can flag patients with elevated mortality risk and CKM burden at no additional cost.

## Introduction

Extracting risk information from images acquired for other indications is an expanding paradigm in cardiovascular medicine. Coronary artery calcium on non-gated chest CT is now recommended for routine reporting by cardiovascular imaging societies and an American Heart Association scientific statement ^1,2^. Such opportunistic markers reach patients who never enter dedicated cardiovascular screening, without additional cost, radiation or acquisition time.

Cardiovascular-kidney-metabolic (CKM) syndrome is a prime target for this approach. It affects most US adults ^3,4^, and the American Heart Association staging framework captures a graded increase in the risk of heart failure and death ^5–7^. However, staging relies on categorical thresholds, and its laboratory criteria are often unavailable in routine care. Substantial heterogeneity also persists within stages ^6,8^. A continuous imaging measure of multisystem burden, derived from scans already performed, could flag patients who warrant CKM evaluation and refine risk within stages.

Bone scintigraphy with technetium-99m-labeled bisphosphonates is among the most frequently performed nuclear imaging examinations, used in oncology, orthopedics and rheumatology ^9^. Since the adoption of nonbiopsy diagnostic criteria for transthyretin cardiac amyloidosis, its use in cardiology has grown and large archives have accumulated ^10^. Most scans show no cardiac uptake (Perugini grade 0) and are read as unremarkable, while the soft tissue signal is dismissed as background. Yet soft tissue tracer retention reflects renal clearance, mineral metabolism and systemic inflammation ^9,11^, each central to CKM pathophysiology. Soft tissue uptake has been linked to amyloid load and mortality in patients with transthyretin amyloidosis ^12^, but whether it tracks CKM burden or carries prognostic information in patients without cardiac uptake is unknown.

We therefore quantified the soft tissue-to-bone ratio (STBR) in 8,769 consecutive patients without cardiac uptake who underwent bone scintigraphy for any indication over a decade. We tested whether STBR is associated with all-cause mortality, major adverse cardiovascular events and heart failure hospitalization independent of established risk factors and CKM stage. We also tested whether renal function or cancer explains this association. Finally, we characterized the clinical traits STBR reflects across 587 diagnoses and measurements and its relationship with CKM stage.

## Methods

### Study Design and Participants

This retrospective single-center cohort study included consecutive patients who underwent ^99m^Tc-DPD whole-body planar bone scintigraphy for any clinical indication at the Vienna General Hospital between January 2010 and August 2020. For patients with multiple scans, the first scan was analyzed. Exclusion criteria were cardiac uptake (Perugini grade ≥1), image quality precluding visual assessment or pathology in both legs precluding measurement (**Supplemental Figure S1**). The ethics committee of the Medical University of Vienna approved the study (2278/2024) and waived informed consent. The study followed the STROBE reporting guideline. Race and ethnicity are not recorded in the institutional records and were therefore not analyzed.

### Data Sources and Outcomes

Demographic, laboratory, echocardiographic and cardiovascular magnetic resonance (CMR) data were extracted from institutional electronic health records. Comorbidities were defined by 3-character ICD-10 codes from medical reports as well as admission and discharge records of the Vienna Health Network, comprising 7 major public hospitals. Vital status was obtained from the national death registry. The primary endpoint was all-cause mortality. Secondary endpoints were major adverse cardiovascular events (MACE; cardiovascular death, nonfatal myocardial infarction or nonfatal stroke) and heart failure hospitalization. Follow-up started at the scan date.

### Image Acquisition and STBR Quantification

Planar anterior and posterior whole-body images were acquired approximately 2.5 hours after intravenous injection of approximately 650 MBq ^99m^Tc-DPD on 1 of 5 dual-head gamma cameras using standardized institutional protocols. Two trained observers blinded to outcomes placed 16-mm circular regions of interest over the mid-diaphyseal femur and tibia of the right leg, each paired with an adjacent soft tissue region. The left leg was used when the right was not assessable (**Supplemental Figures S2 and S3**). STBR was calculated as the ratio of mean soft tissue to mean bone counts per pixel, averaged across the upper and lower leg. Both observers independently read 200 randomly selected scans to assess inter-reader agreement. STBR was analyzed continuously and dichotomized at 0.5.

### Imaging-Phenotype Association Analysis

To characterize the clinical correlates of STBR, we performed an imaging-phenotype association (IPA) analysis analogous to phenome- and genome-wide association studies ^13,14^. Candidate traits comprised 1,160 binary traits (ICD-10 diagnoses, sex, referral source) and 50 continuous clinical measures. Of these, 587 met prespecified inclusion criteria (≥10 individuals per category for binary traits; non-zero variance for continuous traits) and were successfully tested. Each trait was regressed on STBR separately using logistic (binary traits) or linear (continuous traits) regression adjusted for age and cancer status. Bonferroni and Benjamini-Hochberg false discovery rate corrections were applied across all tested traits.

### Cardiovascular-Kidney-Metabolic Staging

Patients were assigned to CKM stages 0 to 4 according to the American Heart Association framework ^5^, adapted to available data (**Supplemental Table S1**). Chronic kidney disease was ascertained by eGFR alone and adiposity by BMI alone. Because CKM-defining variables were frequently missing, 4 analytic strategies were specified before the CKM analysis. These were a best-evidence-available approach as the primary analysis, a complete-case analysis, multiple imputation by chained equations with 20 imputations and the best-evidence-available approach restricted to patients without cancer. The association between STBR and CKM stage was quantified by Spearman correlation, pooled across imputations using Fisher z transformation under Rubin rules.

## Statistical Analysis

Continuous variables are reported as mean (SD) or median (IQR) and were compared using Welch t-tests, Mann-Whitney U tests, Chi-square or Fisher exact tests. Cox proportional hazards models were fitted unadjusted and adjusted for 10 established risk factors, namely age, sex, BMI, renal insufficiency, cancer, diabetes, hypertension, coronary artery disease, atrial fibrillation and liver disease. STBR was modeled dichotomized, per 1-SD increase and with restricted cubic splines and 4 knots. The proportional hazards assumption was assessed using scaled Schoenfeld residuals. Mortality analyses were repeated in subgroups defined by age (<65 vs ≥65 years), sex, BMI (<25 vs ≥25), cancer, diabetes, coronary artery disease, atrial fibrillation, renal insufficiency and cardiology referral. Incremental value of continuous STBR was assessed over 2 base models, including the 10-covariate model and a model of CKM stage, age and sex. We used likelihood ratio tests and the change in Harrell C statistic, with 95% CIs from 500 bootstrap samples. In patients with measured eGFR, a model of age plus STBR was compared with a model of age plus eGFR. Effect modification by CKM stage was tested using an interaction term. Inter-reader agreement was assessed using the intraclass correlation coefficient and Bland-Altman analysis. All tests were two-sided with P < 0.05 considered significant. Analyses were performed in Python version 3.9.5.

## Results

### Patient Characteristics

Of 9,174 patients, 318 were excluded for cardiac uptake, 4 for insufficient image quality and 83 for bilateral lower-limb pathology, yielding 8,769 patients (**Supplemental Figure S1**). Mean (SD) age was 61 (16) years, 3,066 (35.0%) were men, 6,892 (78.6%) had a history of cancer and 982 (11.2%) had bone metastases (**Table 1**). Mean (SD) STBR was 0.35 (0.11). Inter-reader agreement was excellent (intraclass correlation coefficient, 0.93; 95% CI, 0.90-0.95; bias, 0.005; limits of agreement, -0.069 to 0.080; **Supplemental Figure S4**). During a median (IQR) follow-up of 5.1 (2.5-8.2) years, 2,418 patients (27.6%) died.

**Table 1.** Clinical characteristics of the study cohort. Characteristics are shown for the overall cohort and stratified by STBR>0.5. BMI=body mass index; CRP=c-reactive protein; eGFR=estimated glomerular filtration rate; LDL=low density lipoprotein; NA=not available; proBNP=pro-b-type natriuretic peptide; STBR=soft tissue to bone ratio

| **Characteristic** | **All patients<br>(n = 8,769)** | **STBR ≤ 0.5<br>(n = 7,997)** | **STBR > 0.5<br>(n = 772)** | **P value** |
|---|---|---|---|---|
| STBR, mean (SD) | 0.35 (0.11) | 0.33 (0.08) | 0.58 (0.10) | <0.001 |
| Age, years, mean (SD) | 61.2 (15.7) | 60.4 (15.6) | 69.8 (13.5) | <0.001 |
| Sex, No. (%) |  |  |  | <0.001 |
| Women | 5,703 (65.0) | 5,356 (67.0) | 347 (44.9) |  |
| Men | 3,066 (35.0) | 2,641 (33.0) | 425 (55.1) |  |
| BMI, kg/m<sup>2</sup>, mean (SD) | 26.4 (5.3) | 26.4 (5.2) | 26.6 (6.2) | 0.29 |
| Missing | 264 (3.0) | 243 (3.0) | 21 (2.7) |  |
| Cardiology referral, No. (%) | 812 (9.3) | 616 (7.7) | 196 (25.4) | <0.001 |
| Cancer, No. (%) | 6,892 (78.6) | 6,358 (79.5) | 534 (69.2) | <0.001 |
| Bone metastases, No. (%) | 982 (11.2) | 854 (10.7) | 128 (16.6) | <0.001 |
| Renal insufficiency, No. (%) | 706 (8.1) | 543 (6.8) | 163 (21.1) | <0.001 |
| Coronary artery disease, No. (%) | 1,072 (12.2) | 845 (10.6) | 227 (29.4) | <0.001 |
| Diabetes, No. (%) | 1,058 (12.1) | 845 (10.6) | 213 (27.6) | <0.001 |
| Heart failure, No. (%) | 891 (10.2) | 706 (8.8) | 185 (24.0) | <0.001 |
| Amyloidosis, No. (%) | 45 (0.5) | 18 (0.2) | 27 (3.5) | <0.001 |
| Multiple myeloma, No. (%) | 80 (0.9) | 57 (0.7) | 23 (3.0) | <0.001 |
| Monoclonal protein positive, No. (%) | 226 (17.7) | 167 (16.8) | 59 (21.0) | 0.10 |
| Missing | 7,492 (85.4) | 7,001 (87.5) | 491 (63.6) |  |
| NT-proBNP, pg/mL, median (IQR) | 652 (192-2,254) | 495 (147-1,703) | 1,953 (654-5,721) | <0.001 |
| Missing | 7,297 (83.2) | 6,816 (85.2) | 481 (62.3) |  |
| eGFR, mL/min/1.73 m<sup>2</sup>, mean (SD) | 81.0 (31.2) | 82.5 (30.5) | 71.1 (34.1) | <0.001 |
| Missing | 3,964 (45.2) | 3,814 (47.7) | 150 (19.4) |  |
| Total cholesterol, mg/dL, mean (SD) | 184.8 (51.9) | 189.0 (50.2) | 160.1 (55.2) | <0.001 |
| Missing | 5,277 (60.2) | 5,009 (62.6) | 268 (34.7) |  |
| LDL cholesterol, mg/dL, mean (SD) | 106.7 (43.4) | 109.9 (42.4) | 90.7 (44.7) | <0.001 |
| Missing | 6,762 (77.1) | 6,323 (79.1) | 439 (56.9) |  |
| CRP, mg/dL, median (IQR) | 0.68 (0.20-3.36) | 0.59 (0.18-2.80) | 1.79 (0.48-7.51) | <0.001 |
| Missing | 4,311 (49.2) | 4,144 (51.8) | 167 (21.6) |  |
| Hemoglobin, g/dL, mean (SD) | 12.7 (1.9) | 12.8 (1.9) | 11.9 (2.0) | <0.001 |
| Missing | 4,140 (47.2) | 3,978 (49.7) | 162 (21.0) |  |
| Hematocrit, %, mean (SD) | 37.6 (5.4) | 37.8 (5.3) | 35.7 (5.7) | <0.001 |
| Missing | 4,140 (47.2) | 3,978 (49.7) | 162 (21.0) |  |

Patients with STBR greater than 0.5 (n=772; 8.8%) were older (mean [SD] age, 69.8 [13.5] vs 60.4 [15.6] years) and more often men (55.1% vs 33.0%). They also more often had renal insufficiency (21.1% vs 6.8%), heart failure (24.0% vs 8.8%), diabetes (27.6% vs 10.6%) or coronary artery disease (29.4% vs 10.6%) and were more often referred by cardiology (25.4% vs 7.7%) (all P < 0.001). Cancer was less common in this group (69.2% vs 79.5%; P < 0.001). BMI did not differ (mean [SD], 26.6 [6.2] vs 26.4 [5.2]; P = 0.29) (**Table 1**).

### STBR and All-Cause Mortality

STBR greater than 0.5 was associated with all-cause mortality (unadjusted HR, 2.61; 95% CI, 2.34-2.92; adjusted HR [aHR], 1.73; 95% CI, 1.54-1.94; **Figure 1**; **Supplemental Figure S5**). Per 1-SD increase (SD=0.11), the aHR was 1.19 (95% CI, 1.15-1.23). Risk rose in a graded manner across six STBR ranges, with significant differences between all adjacent ranges except the 2 highest ([0.6-0.7] vs >0.7; P=0.60; **Figure 2**). In spline analysis, hazard was stable below the median STBR of 0.34 and increased steeply above it, reaching an aHR of approximately 2 at 0.6 and 3 at 0.80 relative to the median (**Figure 2**). Across thresholds, the aHR increased progressively to 3.42 (95% CI, 2.21-5.32) at STBR greater than 0.76 (**Supplemental Figure S6**).

**Figure 1.**
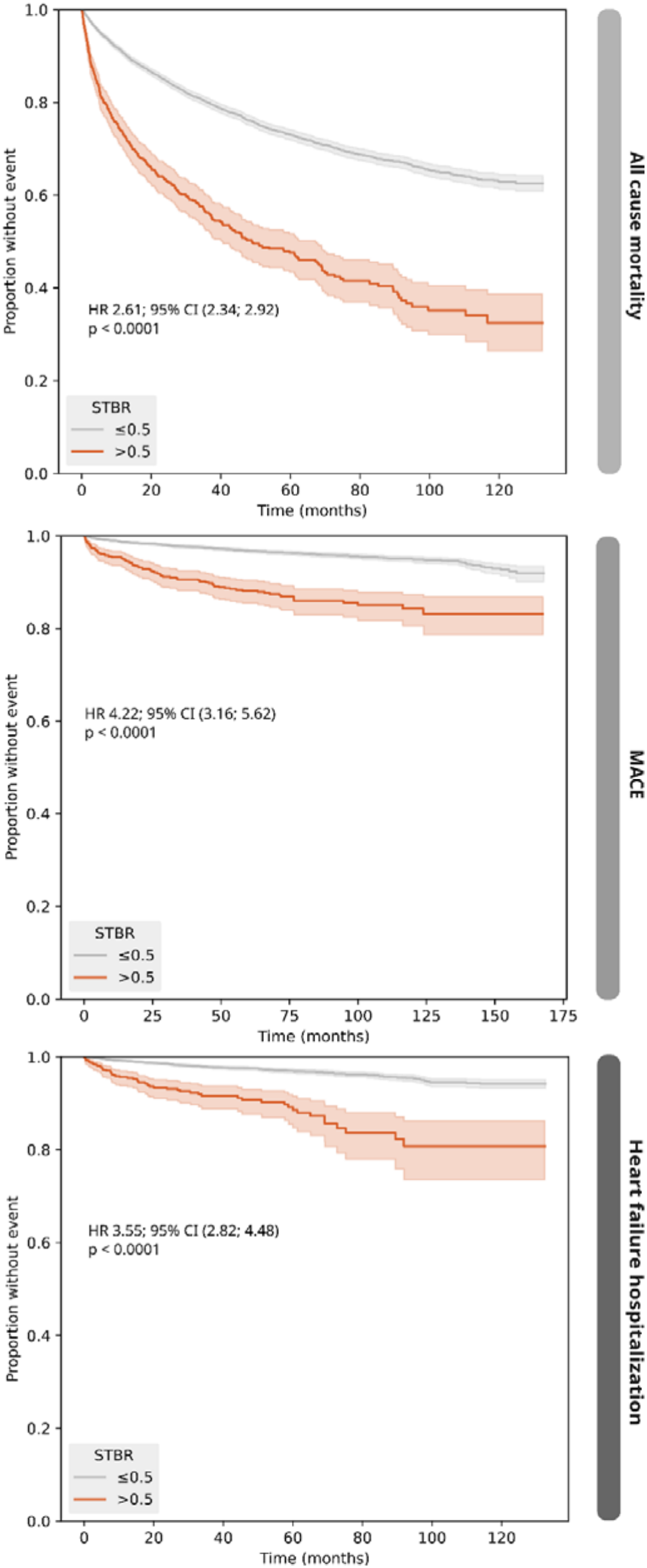
Kaplan-Meier estimators for three endpoints. The estimators show all-cause mortality (top), major adverse cardiovascular event (middle), and heart failure hospitalization (bottom) stratified by STBR>0.5. CI=confidence interval; HR=hazard ratio; MACE=major adverse cardiovascular event

**Figure 2.**
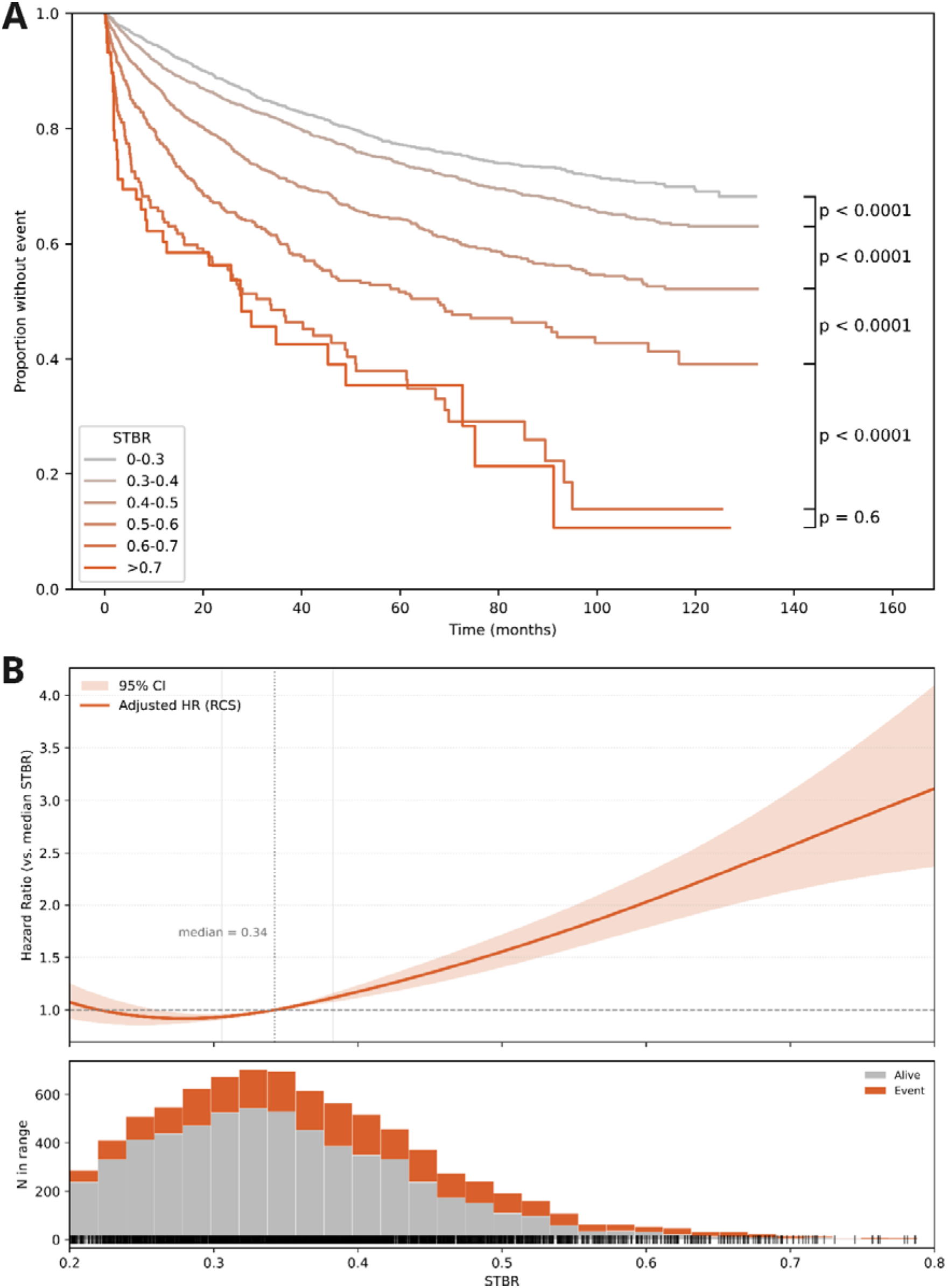
Graded exposure-risk relationship between STBR and all-cause mortality. All-cause mortality for STBR ranges (A) and association between STBR and hazard as restricted cubic spline plot (B) with number of patients per range in each range (bottom panel). Hazard for all-cause mortality increases gradually above the median STBR (0.34) and rises steeply at higher values. Hazard ratios for restricted cubic spline are adjusted for the 10 covariates of the primary model. CI=confidence interval; HR=hazard ratio; STBR=soft tissue to bone ratio

Adding STBR to the 10-covariate model improved fit (likelihood ratio Chi=86.5; P<0.001) and discrimination (C statistic 0.706 to 0.715; ΔC 0.009; 95% CI, 0.007-0.014). In 4,805 patients with measured eGFR, age plus STBR discriminated mortality better than age plus eGFR (ΔC 0.022; 95% CI 0.011-0.032).

### Cardiovascular Endpoints

STBR greater than 0.5 was associated with MACE (unadjusted HR 4.22; 95% CI 3.16-5.62; aHR 1.51; 95% CI 1.11-2.05) and heart failure hospitalization (unadjusted HR 3.55; 95% CI 2.82-4.48; aHR 1.31; 95% CI 1.02-1.67) (**Figure 1**). Per 1-SD increase, aHRs were 1.21 (95% CI 1.10-1.33) for MACE and 1.16 (95% CI 1.06-1.26) for heart failure hospitalization (**Supplemental Figure S7**). Event rates increased in a graded manner across STBR ranges for both endpoints (**Supplemental Figure S8**).

### Independence From Renal Function, Cancer and Other Comorbidities

STBR per SD remained associated with mortality in every prespecified subgroup (all P<0.05; **Supplemental Figure S9**), with aHRs ranging from 1.10 (95% CI 1.02-1.19) in patients with renal insufficiency to 1.45 (95% CI 1.28-1.64) in patients without cancer. The association persisted in patients without renal insufficiency (aHR 1.23; 95% CI 1.18-1.28) and was stronger in patients without cancer (aHR 1.45; 95% CI 1.28-1.64) than in those with cancer (aHR 1.18; 95% CI 1.14-1.22). It was also present in women (aHR 1.25; 95% CI 1.19-1.30), men (aHR 1.13; 95% CI 1.08-1.19), cardiology referrals (aHR 1.36; 95% CI 1.18-1.57) and patients referred for other indications (aHR 1.20; 95% CI 1.15-1.24).

### Imaging-Phenotype Associations

The IPA analysis identified 57 traits significant after Bonferroni correction (39 binary, 18 continuous) and an additional 63 when performing false discovery rate correction (**Figure 3**; **Supplemental Table S2**). The strongest positive binary associations were dominated by CKM-related conditions, including type 2 diabetes, chronic kidney disease, chronic ischemic heart disease, heart failure, atrial fibrillation, peripheral vascular disease and hypertension, followed by liver disease. Breast cancer was the only inverse association, consistent with the higher proportion of men among patients with high STBR. Among continuous measures, STBR was positively associated with CRP, NT-proBNP, atrial diameters, blood urea nitrogen, creatinine, HbA1c, bilirubin, CMR extracellular volume fraction and reduced left ventricular systolic function. It was inversely associated with total and LDL cholesterol, hemoglobin and hematocrit.

**Figure 3.**
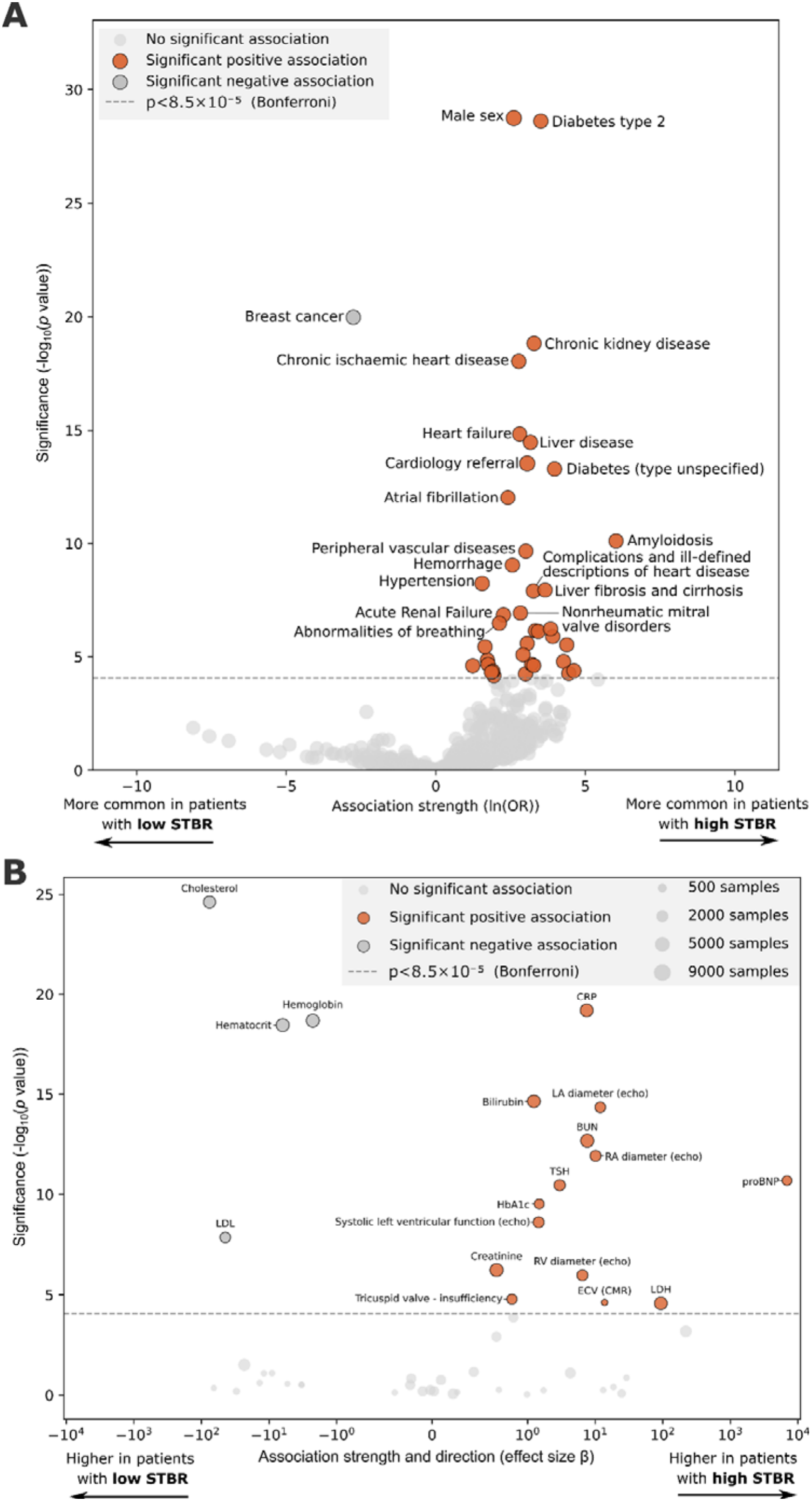
Imaging-phenotype association (IPA) results. Volcano plots show binary traits (A) and continuous traits (B). Left ventricular function is given as dysfunction grades. BMI=body mass index; BUN=blood urea nitrogen; CI=confidence interval; CMR=cardiac magnetic resonance (imaging); CRP=c-reactive protein; ECV=extracellular volume; eGFR=estimated glomerular filtration rate; HbA1c=hemoglobin A1c; HR=hazard ratio; LA=left atrium; ln=natural logarithm; LDH=lactate dehydrogenase; LDL=low density lipoprotein; OR=odds ratio; proBNP=pro-b-type natriuretic peptide; RA=right atrium; RV=right ventricle; STBR=soft tissue to bone ratio; TSH=thyroid stimulating hormone

### Association with CKM Stage

In the primary analysis (n=8,364), mean STBR increased from 0.32 at stage 0 to 0.41 at stage 4 (Spearman ρ=0.30; P<0.001; **Figure 4**). The gradient was consistent in the complete-case analysis (n=1,278; ρ=0.38; P<0.001) and after multiple imputation (n=8,769; ρ=0.25; P <0.001). It was strongest in patients without cancer (n=1,796; ρ=0.40; P<0.001), in whom mean STBR increased from 0.31 to 0.43.

**Figure 4.**
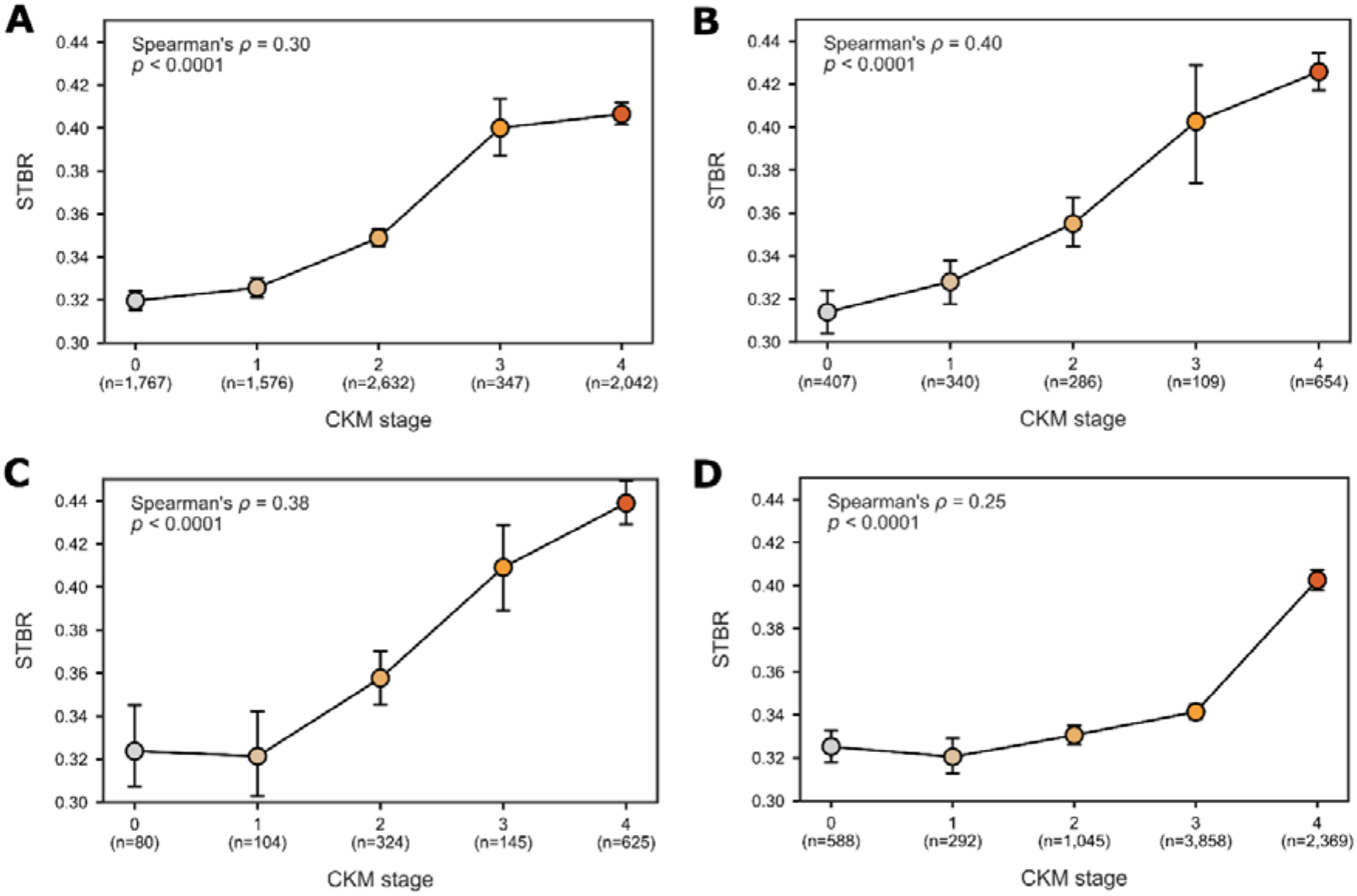
Association of STBR with CKM stage. The results are shown for four separate analyses to ensure robustness: best evidence available among all-comers (A), non-oncological patients (B), complete-case (C), and multiple imputation (D). CKM=cardiovascular-kidney-metabolic (syndrome); STBR=soft tissue to bone ratio

STBR added prognostic information beyond CKM stage, age and sex (likelihood ratio Chi=78.9; P<0.001; ΔC, 0.010; 95% CI, 0.007-0.015). Within each CKM stage, including stage 0, higher STBR was associated with mortality (age- and sex-adjusted HR per SD, 1.19-1.30; all P≤0.004), without evidence of effect modification (P for interaction=0.58).

## Discussion

In 8,769 consecutive patients without cardiac uptake on bone scintigraphy, soft tissue signal routinely dismissed as background was associated with all-cause mortality, MACE and heart failure hospitalization independent of 10 established risk factors. Risk increased in a graded manner with STBR. The association was not explained by renal function, cancer or any other single comorbidity. STBR mapped predominantly to cardiovascular, renal and metabolic traits, increased across CKM stages and added prognostic information beyond CKM stage. These findings identify STBR as an opportunistic marker of systemic cardiovascular risk that can be derived from scans already performed at most hospitals.

Several observations argue against STBR being a surrogate of a known risk factor. Renal clearance is the principal excretion route of bisphosphonate tracers ^9^, yet STBR remained prognostic in patients without renal insufficiency and outperformed eGFR for mortality discrimination when combined with age. The association was substantially stronger in patients without cancer, arguing against STBR being a purely cancer-associated marker. BMI did not differ by STBR category, making soft tissue thickness an unlikely explanation. The association was also consistent across sex, age and referral indication.

The biological basis of STBR is likely multifactorial. Bisphosphonate tracers bind hydroxyapatite-like calcium deposits ^15,16^, and dysregulated mineral metabolism across the CKM spectrum promotes ectopic vascular and soft tissue calcification ^17–19^. The IPA supports a composite signal, with associations spanning cardiac strain (NT-proBNP, chamber dimensions), inflammation (CRP), renal function and hepatic function. Inverse associations with cholesterol and hemoglobin are consistent with the pattern seen in advanced cardiometabolic disease, in which low cholesterol marks frailty rather than favorable risk ^20,21^. Dedicated mechanistic studies are needed to separate these contributions.

These findings are relevant to CKM risk assessment. CKM stage predicts heart failure and death ^6,7^, but staging is categorical and depends on laboratory data that are often missing, as illustrated by unavailable eGFR in 45% of our cohort. STBR increased across stages in all four analytic strategies and stratified mortality within every stage, including stage 0, without effect modification. A continuous imaging index could therefore complement staging by not only identifying patients for CKM evaluation but also for refining risk within stages.

STBR extends the paradigm of opportunistic cardiovascular imaging. Coronary artery calcium on non-gated chest CT predicts cardiovascular and all-cause mortality ^22,23^ and is now recommended for routine reporting ^1,2^. Bone scintigraphy is performed at scale for oncologic, orthopedic and rheumatologic indications as well as increasingly for amyloidosis evaluation ^10,24^. STBR can be annotated in seconds on every routine medical image viewer. Because bone scintigraphy is not part of CKM evaluation, its value lies in opportunistic use. STBR derived from scans acquired for other indications could identify patients with unrecognized CKM burden who may benefit from targeted evaluation of blood pressure, glycemia, lipids, kidney function and albuminuria ^5,25^. In patients with an available scan, STBR may also refine risk within CKM stages and help prioritize intensified management of modifiable risk factors, although whether STBR-guided management improves outcomes requires prospective evaluation. Automated quantification would enable the scalable application to existing archives.

Prior quantitative analyses of bone scintigraphy treated soft tissue activity as a denominator for cardiac uptake rather than as a signal of interest ^26^. In wild-type transthyretin amyloidosis, Tingen et al. linked soft tissue uptake to subcutaneous amyloid load and mortality ^12^. Because these patients typically have advanced age, heart failure, atrial fibrillation and chronic kidney disease, soft tissue uptake in amyloidosis may reflect extracardiac amyloid deposition, comorbid CKM burden or both. Our results show that the soft tissue signal is also prognostic in patients without cardiac uptake but does not distinguish between these mechanisms in patients with amyloidosis.

### Limitations

This study has limitations. First, it was a single-center retrospective study and external validation is required. Second, CKM staging was adapted to available data, using eGFR without albuminuria and BMI without waist circumference. To account for this, we applied four analytic strategies, which yielded concordant gradients. Third, planar imaging is subject to attenuation and overlap, and SPECT could enable volumetric quantification. Fourth, regions of interest were placed manually, although inter-reader agreement was excellent. Fifth, most patients were referred for oncologic indications, reflecting the population undergoing bone scintigraphy. Cancer treatment data were unavailable. Sixth, only ^99m^Tc-DPD was studied. Generalization to other bisphosphonate tracers requires validation.

### Conclusions

Bone scintigraphy has been widely available and routinely performed for decades, yet the soft tissue signal on scans without cardiac uptake has been disregarded as background. In 8,769 such patients, the soft tissue-to-bone ratio was independently associated with all-cause mortality, major adverse cardiovascular events and heart failure hospitalization. The association held in women and men, in younger and older patients and in patients with and without cancer or renal insufficiency. STBR also tracked cumulative cardiovascular-kidney-metabolic burden. Because STBR can be derived from routinely acquired scans without additional imaging, radiation or cost, it may enable opportunistic detection and risk stratification.

## Data Availability

The current ethical approval does not allow sharing of data. All data produced in the present study may be made available upon reasonable request to the authors and a corresponding ethical approval.

## Author contributions

Dr. Spielvogel had full access to all of the data in the study and takes responsibility for the integrity of the data and the accuracy of the data analysis.

Concept and design: Spielvogel

Acquisition, analysis, or interpretation of data: Spielvogel, Kluge, Khanna, Kassem, Ning, Kumpf, Nitsche, Hengstenberg, Slomka, Hacker

Drafting of the manuscript: Spielvogel

Critical review of the manuscript for important intellectual content: All authors

Statistical analysis: Spielvogel

Obtained funding: Spielvogel

Administrative, technical, or material support: Kumpf

Supervision: Hengstenberg, Slomka, Hacker

## Data sharing

Sharing of patient data is not approved under the current IRB approval. For access to data, please contact the corresponding author and/or the Data Clearing House of the Medical University of Vienna.

