## Supplemental Material for "An Overlooked Signal on Routine Bone Scans: Soft Tissue Uptake, Mortality and Cardiovascular-Kidney-Metabolic Burden"

### Supplement

**Table S1.** Operationalization of Cardiovascular-Kidney-Metabolic (CKM) Stages.

| **Stage** | **AHA definition (Ndumele et al, 2023)** | **Operationalization in this study** |
| --- | --- | --- |
| CKM risk feature (used for stages 3 and 4) | Excess adiposity, dysglycemia, metabolic risk factors or CKD | ≥1 of the following: BMI ≥25 kg/m²; HbA1c ≥5.7%; hypertension [ICD-10 I10-I15]; diabetes [ICD-10 E10-E14]; triglycerides ≥135 mg/dL; eGFR <60 mL/min/1.73 m² |
| 4 | Clinical CVD in patients with excess adiposity, metabolic risk factors or CKD | ≥1 CKM risk feature plus ≥1 of the following: heart failure [I50]; atrial fibrillation [I48]; stroke [I63-I64]; coronary artery disease [I20-I25]; myocardial infarction [I21-I22]. Stage 4 was not subdivided into 4a and 4b |
| 3 | Subclinical CVD or risk equivalents (very high-risk CKD, high predicted 10-year CVD risk) | eGFR <30 mL/min/1.73 m², or ≥1 CKM risk feature plus NT-proBNP ≥125 pg/mL or high-sensitivity troponin T ≥22 ng/L in men or ≥14 ng/L in women |
| 2 | Metabolic risk factors (hypertriglyceridemia, hypertension, diabetes, metabolic syndrome) or moderate- to high-risk CKD | Hypertension [ICD-10], diabetes [ICD-10] or HbA1c ≥6.5%, triglycerides ≥135 mg/dL, or eGFR 30-59 mL/min/1.73 m² |
| 1 | Excess or dysfunctional adiposity (BMI ≥25 kg/m² or elevated waist circumference) and/or prediabetes | BMI ≥25 kg/m², or HbA1c 5.7%-6.4% |
| 0 | No CKM risk factors | BMI measured and <25 kg/m², total cholesterol <200 mg/dL or not measured, no CVD, and no criteria for stages 1-4 met |
| Not staged | NA | CVD without any CKM risk feature (n = 115), or criteria for stage 0 not met because BMI was missing or total cholesterol was ≥200 mg/dL (n = 290) |

Stages were assigned hierarchically from stage 4 to stage 0. BMI, body mass index; CKD, chronic kidney disease; CVD, cardiovascular disease; eGFR, estimated glomerular filtration rate; HbA1c, hemoglobin A1c; NA, not applicable; NT-proBNP, N-terminal pro-B-type natriuretic peptide.

**Table S2.** IPA analysis results corrected using Bonferroni and Benjamini-Hochberg FDR correction.

| **Parameter** | **type** | **pval** | **Significant after Bonferroni** | **Significant after BH-FDR** |
| --- | --- | --- | --- | --- |
| Male sex | Binary | <0.0001 | yes | yes |
| Diabetes type 2 | Binary | <0.0001 | yes | yes |
| Cholesterol | Continuous | <0.0001 | yes | yes |
| Breast cancer | Binary | <0.0001 | yes | yes |
| CRP | Continuous | <0.0001 | yes | yes |
| Chronic kidney disease | Binary | <0.0001 | yes | yes |
| Hemoglobin | Continuous | <0.0001 | yes | yes |
| Hematocrit | Continuous | <0.0001 | yes | yes |
| Chronic ischaemic heart disease | Binary | <0.0001 | yes | yes |
| Heart failure | Binary | <0.0001 | yes | yes |
| Bilirubin | Continuous | <0.0001 | yes | yes |
| Liver disease | Binary | <0.0001 | yes | yes |
| LA diameter (echo) | Continuous | <0.0001 | yes | yes |
| Cardiology referral | Binary | <0.0001 | yes | yes |
| Diabetes (type unspecified) | Binary | <0.0001 | yes | yes |
| BUN | Continuous | <0.0001 | yes | yes |
| Atrial fibrillation | Binary | <0.0001 | yes | yes |
| RA diameter (echo) | Continuous | <0.0001 | yes | yes |
| proBNP | Continuous | <0.0001 | yes | yes |
| TSH | Continuous | <0.0001 | yes | yes |
| Amyloidosis | Binary | <0.0001 | yes | yes |
| Peripheral vascular diseases | Binary | <0.0001 | yes | yes |
| HbA1c | Continuous | <0.0001 | yes | yes |
| Hemorrhage | Binary | <0.0001 | yes | yes |
| Systolic left ventricular function (echo) | Continuous | <0.0001 | yes | yes |
| Hypertension | Binary | <0.0001 | yes | yes |
| Fibrosis and cirrhosis of the liver | Binary | <0.0001 | yes | yes |
| Complications and ill-defined descriptions of heart disease | Binary | <0.0001 | yes | yes |
| LDL | Continuous | <0.0001 | yes | yes |
| Nonrheumatic mitral valve disorders | Binary | <0.0001 | yes | yes |
| Acute Renal Failure | Binary | <0.0001 | yes | yes |
| Abnormalities of breathing | Binary | <0.0001 | yes | yes |
| Other diseases of biliary tract | Binary | <0.0001 | yes | yes |
| Creatinine | Continuous | <0.0001 | yes | yes |
| Erysipelas | Binary | <0.0001 | yes | yes |
| Diseases of stomach and duodenum | Binary | <0.0001 | yes | yes |
| RV diameter (echo) | Continuous | <0.0001 | yes | yes |
| Alcoholic liver disease | Binary | <0.0001 | yes | yes |
| Respiratory failure | Binary | <0.0001 | yes | yes |
| Polyneuropathy | Binary | <0.0001 | yes | yes |
| Other COPD | Binary | <0.0001 | yes | yes |
| Transplanted organ or tissue | Binary | <0.0001 | yes | yes |
| Other anemias | Binary | <0.0001 | yes | yes |
| Retinal disorders in diseases classified elsewhere | Binary | <0.0001 | yes | yes |
| Tricuspid valve - insufficiency | Continuous | <0.0001 | yes | yes |
| Mental and behavioural disorders due to use of alcohol | Binary | <0.0001 | yes | yes |
| Stroke, not specified as haemorrhage or infarction | Binary | <0.0001 | yes | yes |
| Fracture of rib(s), sternum and thoracic spine | Binary | <0.0001 | yes | yes |
| ECV (CMR) | Continuous | <0.0001 | yes | yes |
| Disorders of lipoprotein metabolism and other lipidaemias | Binary | <0.0001 | yes | yes |
| LDH | Continuous | <0.0001 | yes | yes |
| Injury of unspecified body region | Binary | <0.0001 | yes | yes |
| Pneumonia, organism unspecified | Binary | <0.0001 | yes | yes |
| Presence of cardiac and vascular implants and grafts | Binary | <0.0001 | yes | yes |
| Decubitus ulcer and pressure area | Binary | 5.41E-05 | yes | yes |
| Multiple myeloma and malignant plasma cell neoplasms | Binary | 5.68E-05 | yes | yes |
| Other disorders of fluid, electrolyte and acid-base balance | Binary | 6.75E-05 | yes | yes |
| Oesophageal varices | Binary | 8.72E-05 | no | yes |
| Systemic sclerosis | Binary | 0.000101 | no | yes |
| Ulcer of lower limb, not elsewhere classified | Binary | 0.000115 | no | yes |
| Personal history of medical treatment | Binary | 0.000119 | no | yes |
| Atherosclerosis | Binary | 0.00013 | no | yes |
| Acute myocardial infarction | Binary | 0.000134 | no | yes |
| Diastolic left ventricular function (echo) | Continuous | 0.000137 | no | yes |
| Haemorrhage from respiratory passages | Binary | 0.000141 | no | yes |
| Bacterial pneumonia, not elsewhere classified | Binary | 0.000187 | no | yes |
| Iron deficiency anaemia | Binary | 0.000281 | no | yes |
| Toxic liver disease | Binary | 0.000283 | no | yes |
| Other neoplasms of uncertain or unknown behaviour of lymphoid, haematopoietic and related tissue | Binary | 0.000484 | no | yes |
| Type 1 diabetes mellitus | Binary | 0.000508 | no | yes |
| Creatine kinase | Continuous | 0.000655 | no | yes |
| Ascites | Binary | 0.000696 | no | yes |
| Elevated blood glucose level | Binary | 0.000697 | no | yes |
| Malignant neoplasm of liver and intrahepatic bile ducts | Binary | 0.000796 | no | yes |
| Other diseases of anus and rectum | Binary | 0.0008 | no | yes |
| Abnormalities of heart beat | Binary | 0.000822 | no | yes |
| Mitral valve - insufficiency | Continuous | 0.001232 | no | yes |
| Other diseases of intestine | Binary | 0.001292 | no | yes |
| Chronic viral hepatitis | Binary | 0.001329 | no | yes |
| Oedema, not elsewhere classified | Binary | 0.001616 | no | yes |
| Acute posthaemorrhagic anaemia | Binary | 0.001629 | no | yes |
| Other dermatitis | Binary | 0.001765 | no | yes |
| Malaise and fatigue | Binary | 0.001831 | no | yes |
| Paraplegia and tetraplegia | Binary | 0.00214 | no | yes |
| Nonrheumatic aortic valve disorders | Binary | 0.002399 | no | yes |
| Gonarthrosis [arthrosis of knee] | Binary | 0.002687 | no | yes |
| Isolated proteinuria | Binary | 0.002742 | no | yes |
| Vasomotor and allergic rhinitis | Binary | 0.002768 | no | yes |
| Unspecified kidney failure | Binary | 0.003176 | no | yes |
| Other disorders of urinary system | Binary | 0.003322 | no | yes |
| Chronic hepatitis, not elsewhere classified | Binary | 0.003348 | no | yes |
| Other polyneuropathies | Binary | 0.003782 | no | yes |
| Superficial injury of head | Binary | 0.004272 | no | yes |
| Other pulmonary heart diseases | Binary | 0.004337 | no | yes |
| Complications of procedures, not elsewhere classified | Binary | 0.004407 | no | yes |
| Nonrheumatic tricuspid valve disorders | Binary | 0.004503 | no | yes |
| Rheumatic tricuspid valve diseases | Binary | 0.004505 | no | yes |
| Other sepsis | Binary | 0.004607 | no | yes |
| Hyperplasia of prostate | Binary | 0.004688 | no | yes |
| Other disorders of veins | Binary | 0.004975 | no | yes |
| Unspecified haematuria | Binary | 0.005391 | no | yes |
| Acute bronchitis | Binary | 0.005445 | no | yes |
| Glomerular disorders in diseases classified elsewhere | Binary | 0.006063 | no | yes |
| Gangrene, not elsewhere classified | Binary | 0.006165 | no | yes |
| Aortic aneurysm and dissection | Binary | 0.006419 | no | yes |
| Delirium, not induced by alcohol and other psychoactive substances | Binary | 0.006491 | no | yes |
| Symptoms and signs concerning food and fluid intake | Binary | 0.006527 | no | yes |
| Hepatic failure, not elsewhere classified | Binary | 0.006554 | no | yes |
| Cardiac arrest | Binary | 0.006609 | no | yes |
| Other specified diabetes mellitus | Binary | 0.007738 | no | yes |
| Atrioventricular and left bundle-branch block | Binary | 0.007894 | no | yes |
| Pleural effusion, not elsewhere classified | Binary | 0.00822 | no | yes |
| Volume depletion | Binary | 0.00879 | no | yes |
| Malignant melanoma of skin | Binary | 0.009302 | no | yes |
| Peritonitis | Binary | 0.00947 | no | yes |
| Malignant neoplasm of kidney, except renal pelvis | Binary | 0.009677 | no | yes |
| Other specified bacterial agents as the cause of diseases classified to other chapters | Binary | 0.009982 | no | yes |
| Gastritis and duodenitis | Binary | 0.010315 | no | yes |
| Other disorders involving the immune mechanism, not elsewhere classified | Binary | 0.010346 | no | yes |
| Gastric ulcer | Binary | 0.010357 | no | yes |

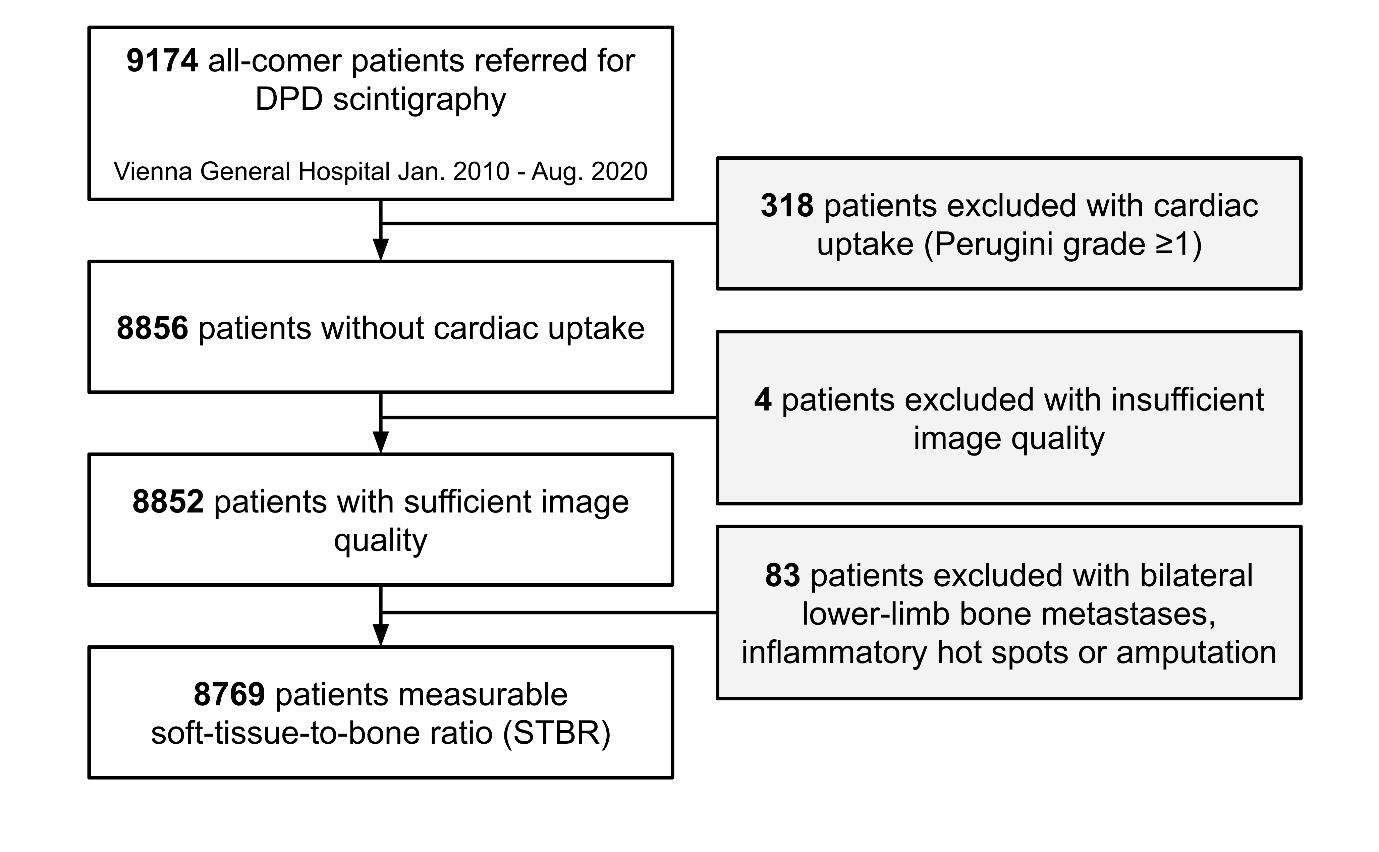

**Figure S1.** Cohort flow diagram. Of 9,174 all-comer patients referred for whole body DPD bone scintigraphy, 8,769 patients were included in the final analysis.

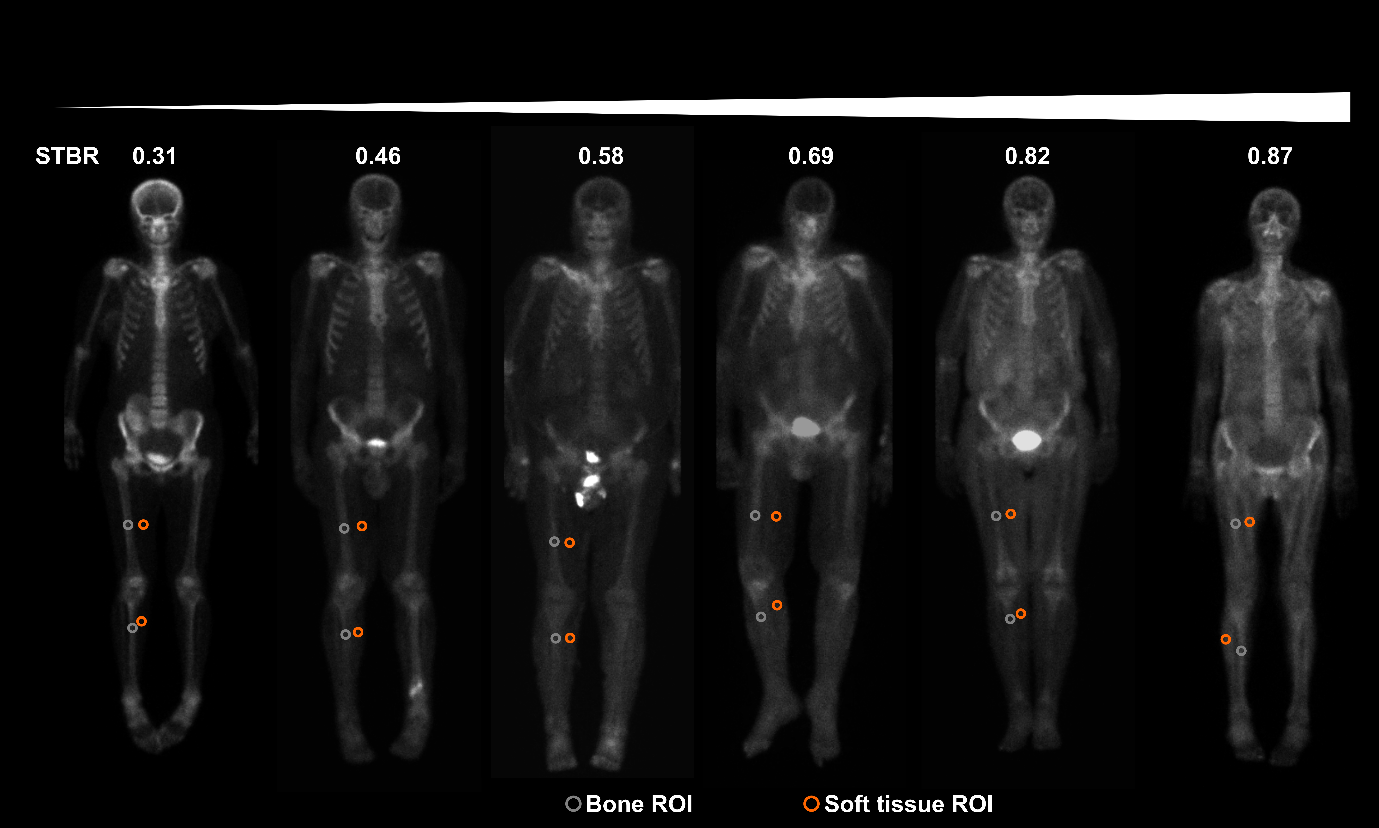

**Figure S2.** Examples of annotated bone scintigraphy images.

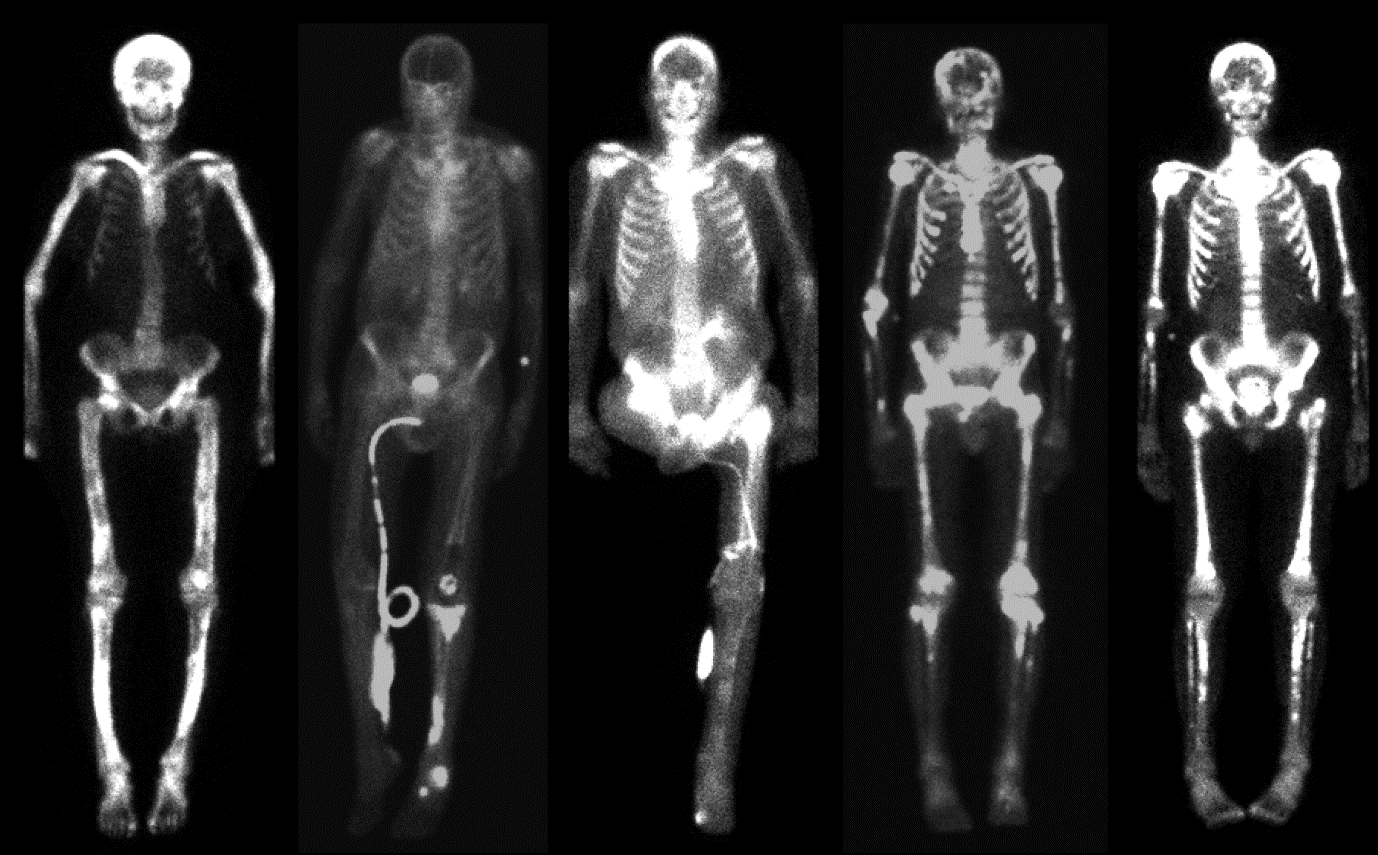

**Figure S3.** Examples of excluded patients due to pathological uptake or missing extremities.

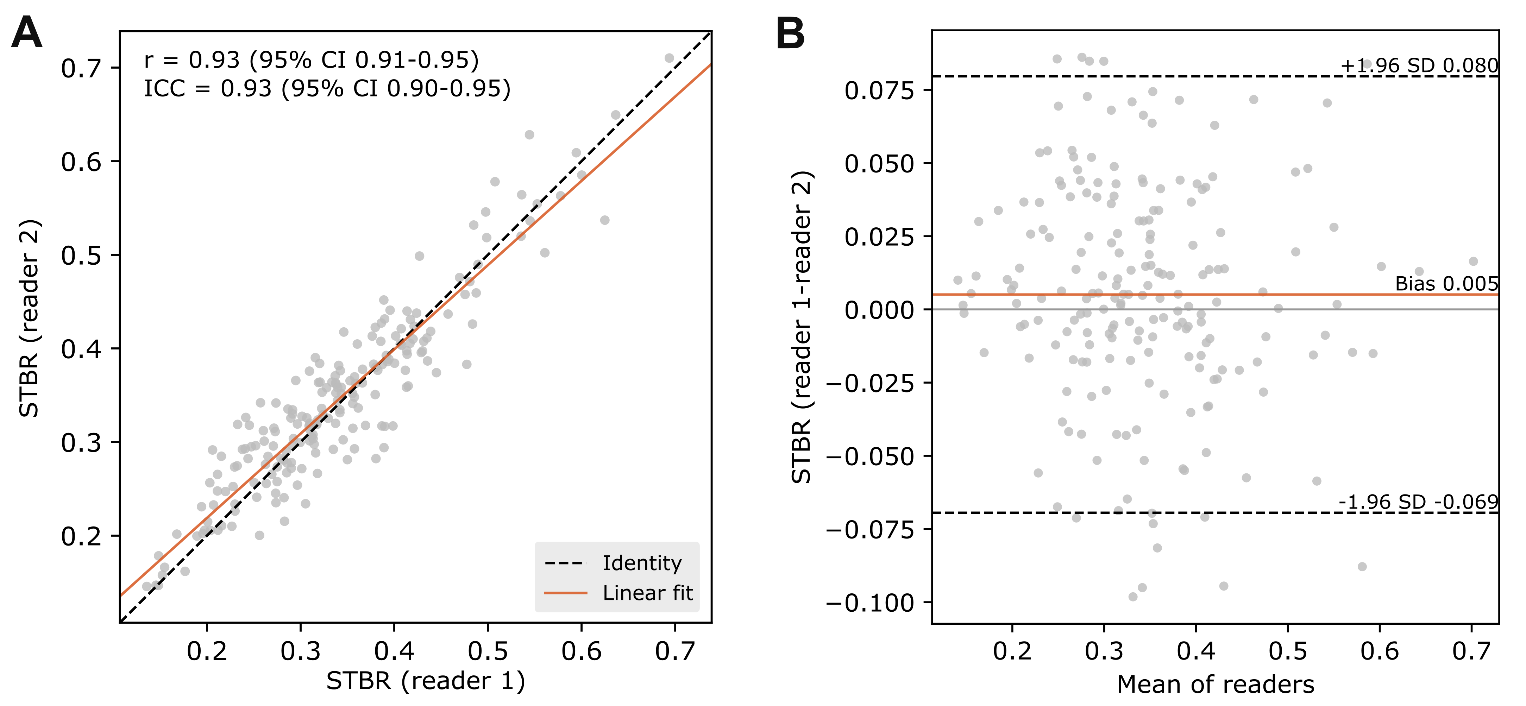

**Figure S4.** Inter-reader agreement of STBR in 200 scans quantified independently by two blinded readers. (A) Reader 2 vs reader 1 with identity line and linear fit. (B) Bland-Altman plot showing mean bias and 95% limits of agreement (±1.96 SD).

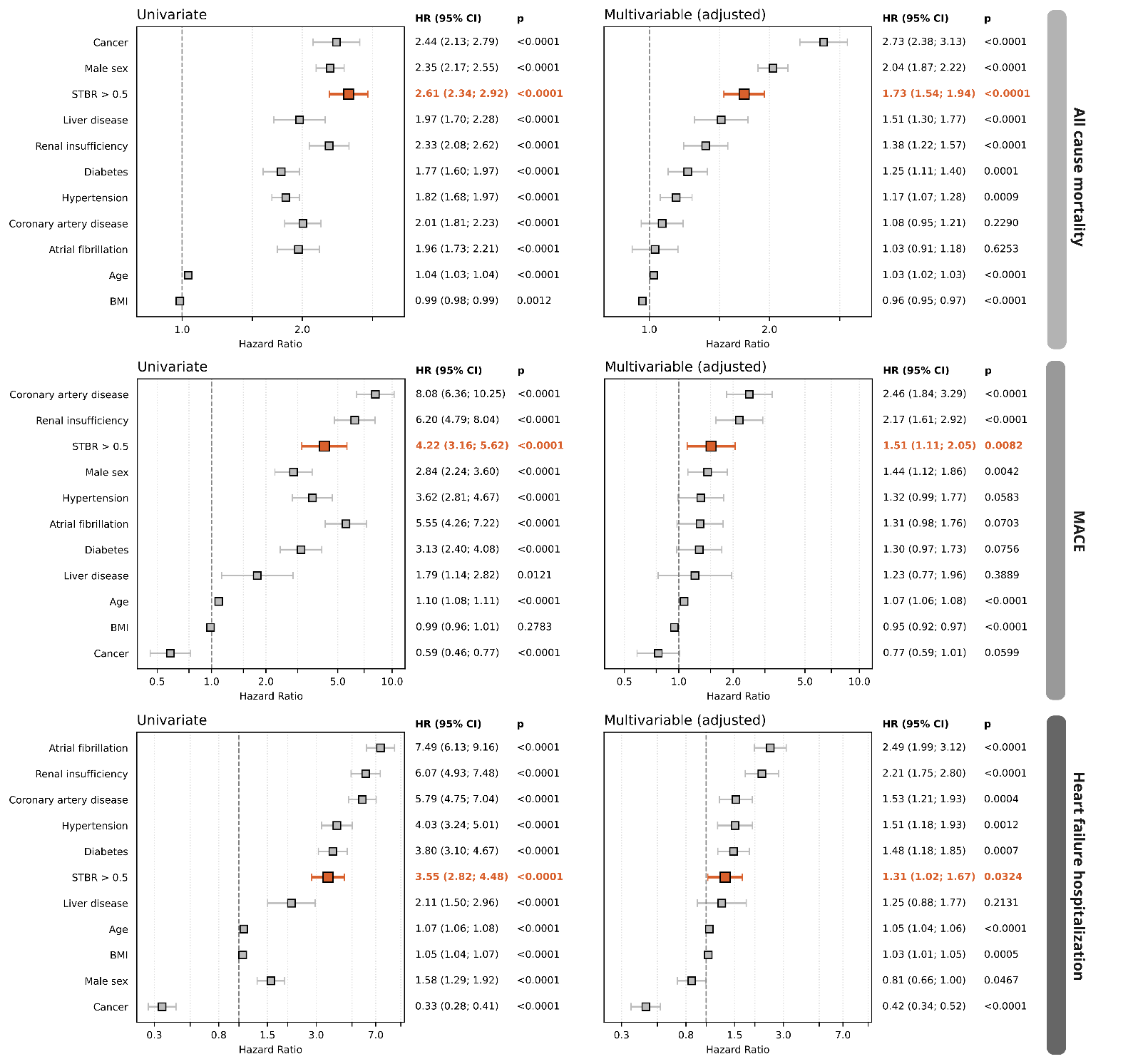

**Figure S5. Association of STBR with outcomes with thresholded STBR.** Hazard ratios of univariable (left) and adjusted multivariable (right) Cox proportional hazards models of STBR thresholded at 0.5 for all-cause mortality (top row), MACE (middle row), and heart failure hospitalization (bottom row). BMI=body mass index; CI=confidence interval; HR=hazard ratio; STBR=soft tissue to bone ratio

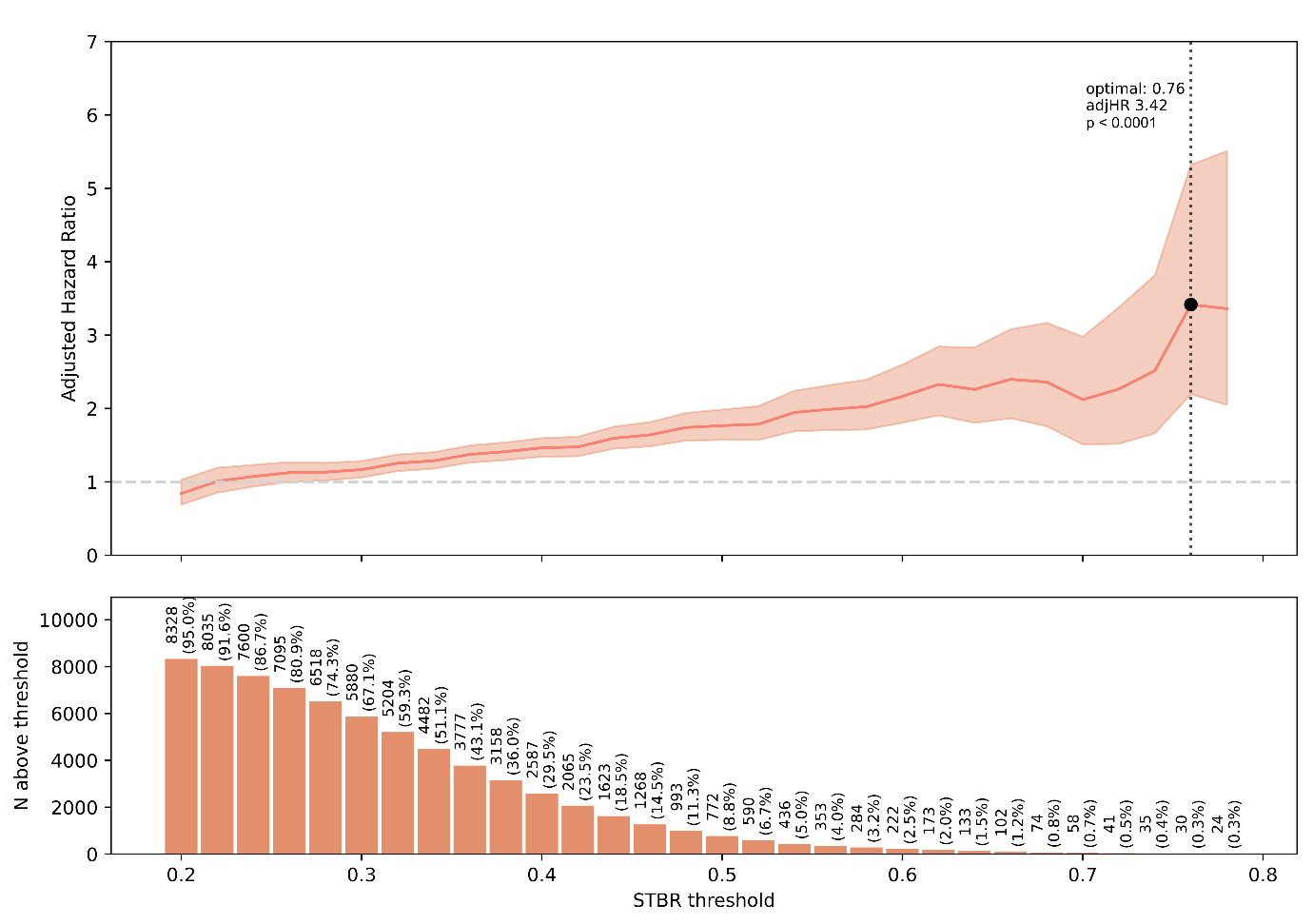

**Figure S6.** All-cause mortality hazard ratios at various STBR thresholds (upper panel) and number of patients above each corresponding threshold (lower panel), adjusted for the same ten covariates as the primary model.

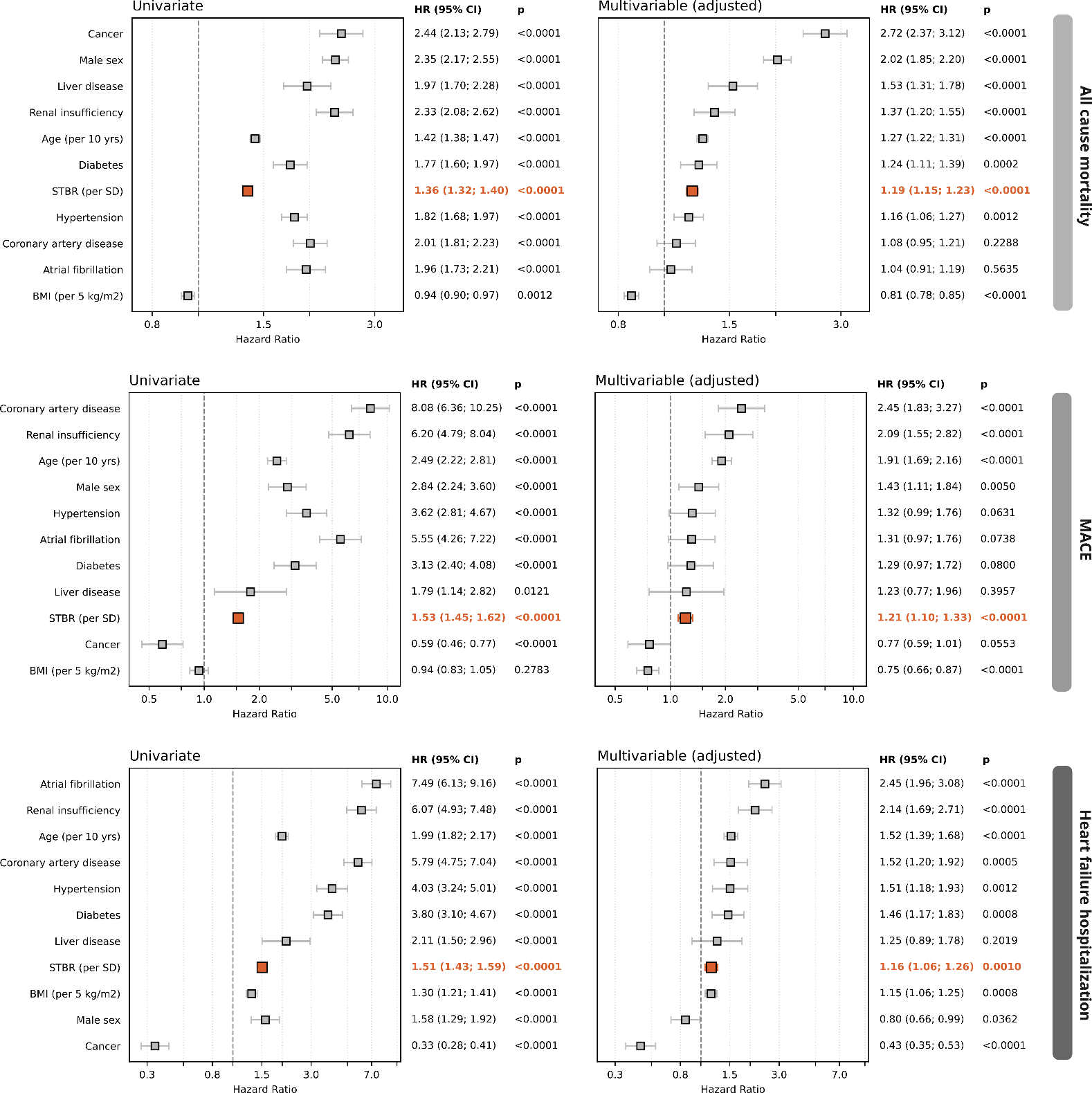

**Figure S7. Association of STBR with outcomes for continuous STBR.** Hazard ratios of univariable (left) and adjusted multivariable (right) Cox proportional hazards models of continuous STBR (per standard deviation) for all-cause mortality (top row), MACE (middle row) and heart failure hospitalization (bottom row).

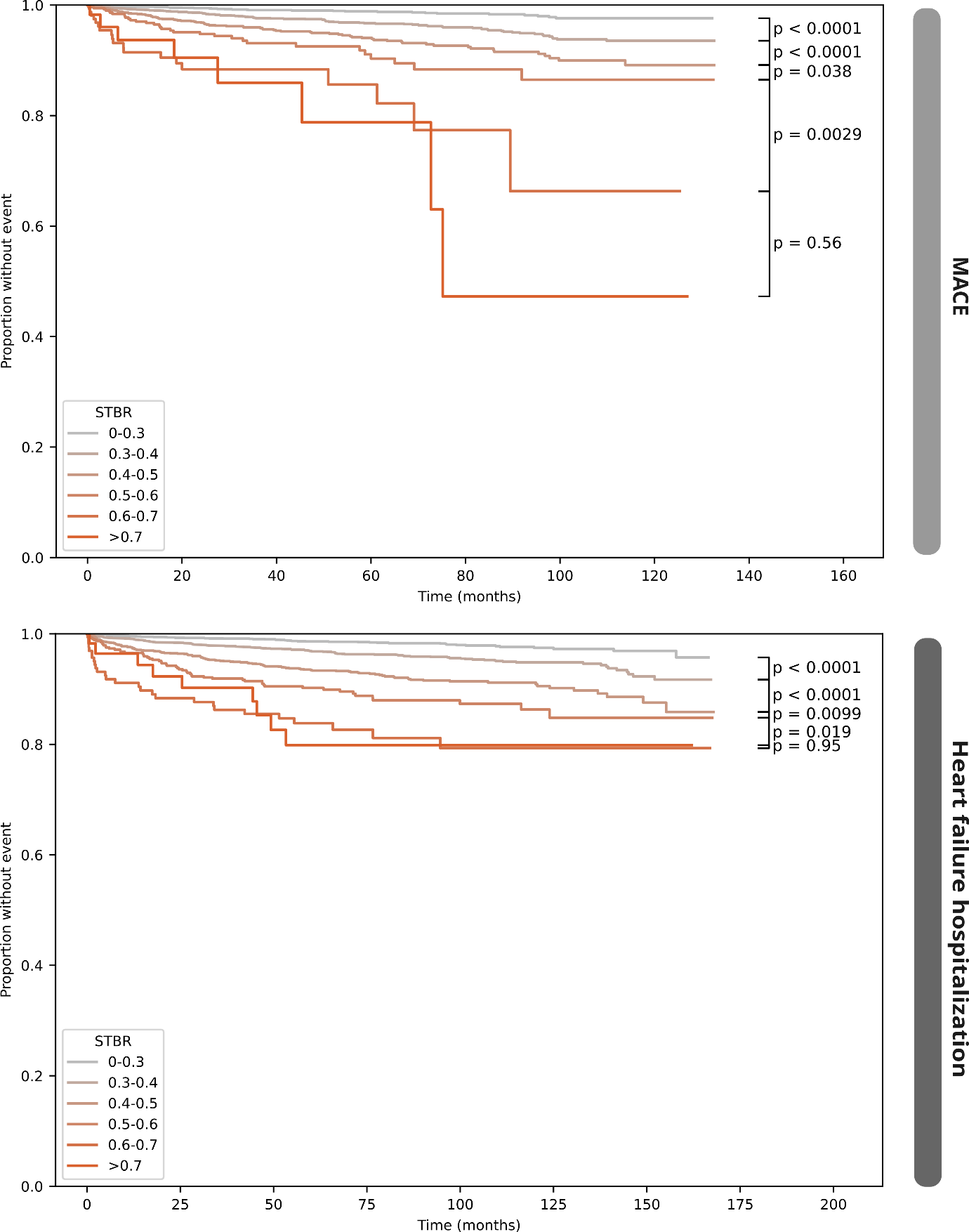

**Figure S8.** Time to event analyses for STBR ranges with MACE (top) and heart failure hospitalization (bottom) as endpoints. Increasing STBR values were associated with higher event rates.

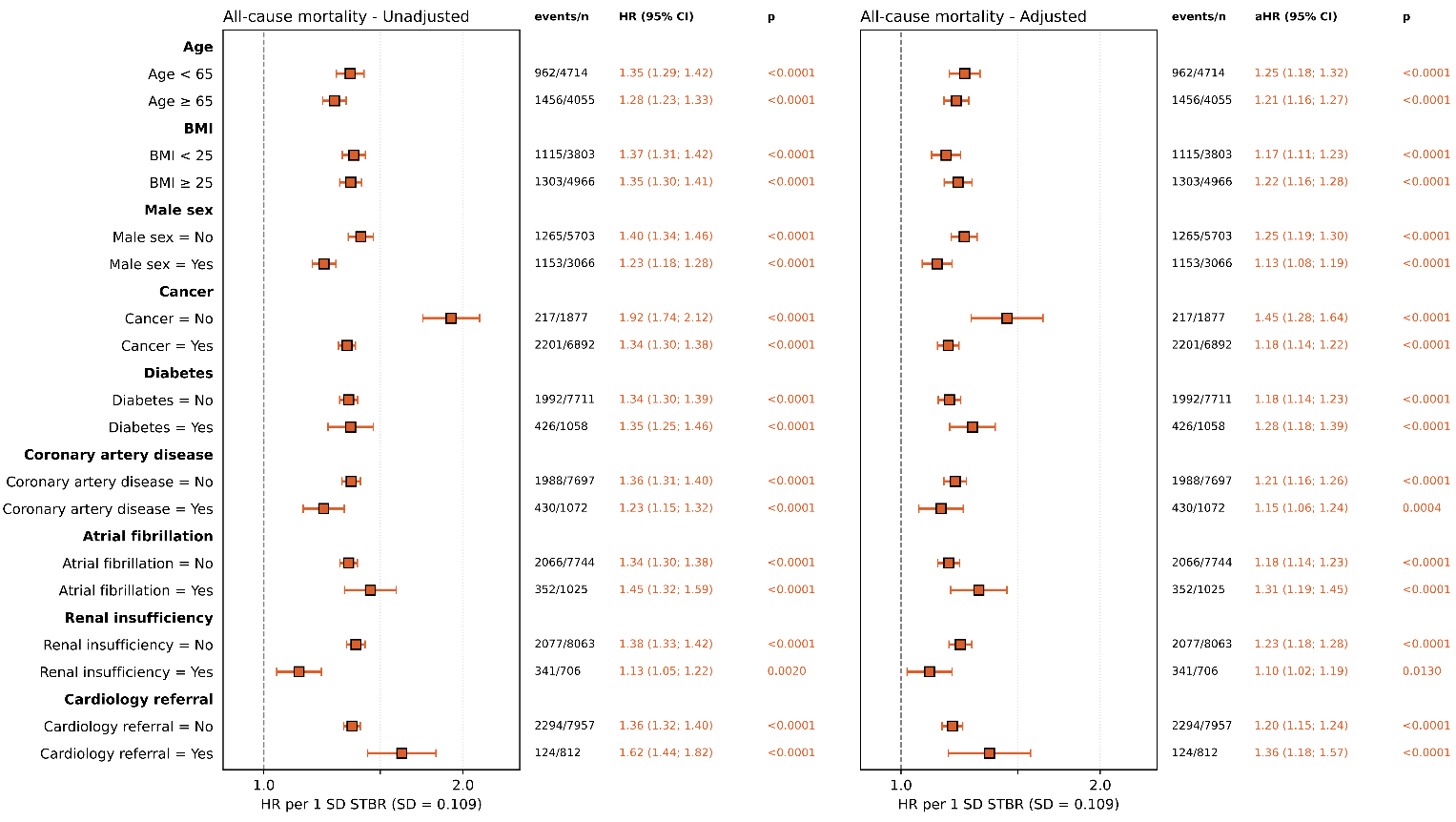

**Figure S9.** Prognostic subgroup and sensitivity analyses for continuous STBR (per SD based on the entire cohort) for all-cause mortality. For each subgroup (row), STBR was corrected for the same parameters as in the primary adjusted model (BMI, age, cancer, sex, renal insufficiency, hypertension, diabetes, coronary artery disease, atrial fibrillation and liver disease). The stratifying variable was left out of the adjustment within each subgroup.
